# Early enteral nutrition after Paediatric Ostomy Closure (EPOC): a protocol for a multicentre, prospective randomised controlled trial

**DOI:** 10.64898/2025.12.24.25342985

**Authors:** Joyce Ly, James Cope, Nicholas Olsen, Soundappan SV Soundappan, Susan Adams

## Abstract

**Introduction:** This is the study protocol of an ongoing prospective randomised controlled trial (RCT) written as per the SPIRIT guidelines. This RCT is designed to assess the safety and efficacy of early enteral nutrition after elective enterostomy closure in paediatric patients.

**Methods and analysis:** This is a multicentre, RCT that will be conducted in two Australian tertiary paediatric hospitals with a planned sample size of 68. Children that meet the inclusion criteria aged between three months and 16 years (inclusive) undergoing an elective enterostomy closure will be invited to participate. To assign treatment group, stratified permutated block randomisation will be used, with block size of four and six, and strata including: age group (3 months - <6 years, 6 - ≤16 years); stoma type (ileostomy, colostomy); and hospital (Sydney Children’s Hospital, Randwick, The Children’s Hospital at Westmead). The control group will commence feeds on return of bowel function or when nasogastric tube aspirates are non-bilious. The intervention group will be offered feeds within 24 hours of admission to the recovery room. The primary outcome is the length of stay, measured in hours. Secondary outcomes include time to oral feeds, time to first spontaneous passage of stool, highest post-operative pain score, complications, analgesia use and parent satisfaction. Data analyses will use an intention-to-treat principle. Length of stay, time to feeds and passing of stool will be analysed using restricted mean survival time. Other secondary outcomes will be analysed using regression models.

**Ethics and dissemination:** The Sydney Children’s Hospital Network Human Research Ethics Committee approved the research (2021/ETH01062). The aim is to report the results in peer-reviewed journals, seminars and conference presentations.

**Trial registration:** ACTRN12621001414808; Australian New Zealand Clinical Trials Registry.

**STRENGTHS AND LIMITATIONS OF THIS STUDY:** *Strengths:*

- This is a stratified, well-powered prospective, randomised controlled trial that minimises heterogeneity between cases with strict eligibility criteria.
- Results will be analysed according to the intention-to-treat principle, utilising statistical methods to fully account for the spectrum of missingness.
- This study includes two separate tertiary paediatric hospitals in Australia, increasing external validity.

*Limitations:*

- Due to the nature of the intervention (timing of feeding), it is not possible to blind participants and researchers to the study group.
- Some outcomes require parent self-reporting, introducing potential bias.

## INTRODUCTION

Reversible enterostomies are commonly indicated in children who have had Hirschsprung’s disease, anorectal malformations and necrotising enterocolitis managed in the first months of life. They are generally located in the colon (colostomy), ileum (ileostomy) and jejunum (jejunostomy) [1]. To close the stoma and restore normal bowel continuity and function, intestinal anastomosis is required. The procedure is relatively low risk but possible post- operative complications include surgical site infection, ileus, bowel obstruction and anastomotic leak [2–6]. Following intestinal anastomosis, patients are traditionally kept nil by mouth (NBM) for a few days until bowel function returns, indicated by the passage of stool or flatus [1]. However, prolonged post-operative fasting may not be necessary. Currently, it is thought that effective anastomotic healing may depend on factors other than feeding, including surgical technique, splanchnic blood flow, preoperative, intraoperative and post- operative management [1, 7]. Indeed, there is evidence that enteral feeding may enhance healing [8].

Early enteral nutrition (EEN) is a core principle of enhanced recovery after surgery (ERAS) and fast-track protocols, which are designed to optimise post-operative recovery through pre- operative, peri-operative and post-operative care [9]. In adult populations, the safety and efficacy of EEN have been extensively documented, with systematic reviews showing significantly shortened length of stay (LOS) [10], reduction in post-operative complications and a decreasing trend in anastomotic leakage and overall mortality [11]. Animal studies have demonstrated increased anastomotic healing and wound strength with EEN [12–15].

There is also increasing evidence that EEN is safe and effective in children. Currently, there have been several randomised controlled trials (RCTs) [2, 3, 16–20], controlled trials [6, 21, 22] and observational studies [4, 23–25] that have shown improved recovery rates, shorter time to full feeds and stool passage, and no negative outcomes associated with EEN. However, existing studies are limited by broad selection criteria and heterogenous definitions of feeding. In addition, it is unclear how EEN impacts pain scores, analgesia use, quality of life or parent satisfaction. Published studies have encouraged further evaluation of the safety and efficacy of EEN in paediatrics using higher quality RCTs with a clearer selection criteria and detailed feeding regime [26].

### Study objectives

This study aims to determine the effect of EEN on recovery after elective enterostomy closure in children. It is hypothesised that post-operative EEN leads to shorter LOS, reduced pain, fewer surgical complications and improved parent satisfaction. The primary objective is to determine whether EEN significantly reduces LOS in paediatric patients after enterostomy closure. The secondary objectives are to assess the effect of EEN on:

1. Time to full feeds and time to pass stool.
2. Post-operative complications.
3. Pain scores and analgesia use.
4. Parent/carer satisfaction at discharge.

## METHODS AND ANALYSIS

### Trial design

This study is designed as a multicentre prospective, open-label RCT with two groups: intervention (EEN) and control (standard practice).

### Study setting

This ongoing study is being conducted at two tertiary hospitals, both of which are in the Sydney Children’s Hospital Network (SCHN): The Children’s Hospital at Westmead (CHW) and Sydney Children’s Hospital, Randwick (SCH). Approximately 40 enterostomy closures occur annually across the two hospitals.

### Inclusion criteria

Paediatric patients between the ages of three months and 16 years (inclusive) will be included if they can likely be fed orally or via nasogastric tube (NGT) after an elective enterostomy closure with the following indications:

1. Elective closure of colostomy after anorectal malformation repair.
2. Elective closure of colostomy/ileostomy after pull-through for Hirschsprung’s disease.
3. Elective closure of ileostomy after previous neonatal laparotomy for necrotising enterocolitis, perforation, atresia or other intestinal pathology that has since been treated or resolved.
4. Elective closure of colostomy/ileostomy of a stoma that was formed from a previous laparotomy for other intraabdominal pathology that has since been treated or resolved.

### Exclusion criteria

Children will be excluded from the study if any of the following are present:

1. Inability to obtain consent from parent/guardian.
2. Patients undergoing a total colectomy or gastrostomy closure.
3. Patients with short gut needing long term total parenteral nutrition.
4. Concomitant conditions likely to affect bowel function (i.e. neurological impairment).
5. Patients evaluated to have laryngeal penetration or unsafe swallow with associated reflux and vomiting. This includes those with structural abnormalities or neurological impairment, due to risk of aspiration.
6. Patients requiring specific personalised feeding regime (i.e. metabolic disease).
7. Patients who require extended NGT feeding or gastrostomy feeding prior to and after surgery that do not have normal gut function.
8. Patients requiring more than a simple enterostomy closure (i.e. extensive adhesiolysis or more than one anastomosis in necrotising enterocolitis).
9. Patients with inflammatory bowel disease or cystic fibrosis.

### Patient and public involvement

Patients and the public will not be involved in the design, administration, reporting or dissemination of this study.

### Recruitment and retention

Upcoming elective operations will be assessed by the investigators for potential participants. Parents/guardians of eligible patients will be contacted by a member of the study team. Where possible, recruitment will be conducted by an investigator that is not part of the patient’s primary treating team. The study will be explained to the parent/guardian and the information sheet provided via email (Online Supplementary Appendix 1). Notably, the sheet contains assurance that a decision not to participate will not affect the standard of their medical care, with the post-operative feeding regime determined instead by their treating surgeon. Contact numbers for the ethics committee in the case that any concerns arise will also be provided. Written consent will be obtained by the investigator on admission to hospital for those that wish to participate (Online Supplementary Appendix 2). Those that consent will then be randomised into treatment groups at the completion of the procedure, once the child arrives in recovery.

Parents/carers may withdraw their consent at any time, and data pertaining to their child will be excluded from analysis. Recruitment will continue until the desired sample size, excluding withdrawals, is reached.

### Allocation

Computer-generated stratified permutated block randomisation will be prepared by the biostatistician. Patients will be stratified by: type of stoma closure (ileostomy or colostomy closure); hospital type (SCH, CHW); and age (3 months – <6 years; or 6 years – ≤16 years), with a block size of four and six. Randomisations will be placed in sequenced sealed opaque envelopes prior to the study by an investigator not involved in the data collection.

At the completion of the procedure and on admission to the recovery room, the patient’s family, the child if old enough, and treating surgical team will be informed of the allocation. Thus, the allocation is not known at the time of surgery. No blinding is possible after this due to the nature of the intervention.

The investigator surgeons will have broad knowledge of previous treatment allocations but not down to the individual blocks. We recognise that this may introduce bias. However, this is mitigated by the operating surgeon not being aware of previous treatment allocations, and the allocation not being revealed until after the operation is completed.

### Intervention group (EEN)

Oral feeds will be offered within 24 hours of admission to the recovery room as tolerated. Unless required for feeding, the NGT will be removed in theatre or in recovery. The first feed will be clear fluids, graded up to full fluids and solids *ad lib*, parent/carer/patient led, as tolerated. Solids will be commenced within 48 hours of admission to the recovery room *ad lib*, parent/carer/patient led as tolerated. If vomiting occurs, the patient will be clinically assessed, fasted for four hours and feeds will be reintroduced, starting with clear fluids, unless clinically contraindicated. Clinical concerns will be addressed as per usual care.

### Control group (standard practice)

The NGT will remain in place post-operatively. Patients will be kept NBM and feeds will be commenced once bowel function returns, as indicated by the passage of flatus or stool, or when NGT aspirates become non-bilious. For those not fed via NGT, it will be removed as per the treating team. The first feed will be clear fluids, gradually graded up to full fluids and solids, as directed by the treating team. If vomiting occurs, the patient will be fasted for four hours and feeds reintroduced starting with clear fluids, unless clinically contraindicated.

### Outcomes

The primary outcome for this study is the treatment efficacy of EEN compared to standard practice management following enterostomy closure, measured as post-operative LOS. This is defined as from time of surgery stopping to discharge in hours. Discharge time will be taken from the electronic medical record, representing the time patients leave the hospital.

Secondary outcomes are:

1. Time to first oral feeds in hours, including clear fluids, full fluids and full enteral intake.
2. Time to first spontaneous passage of stool in hours.
3. Highest post-operative pain score, measured at any time during the hospital stay, using the FLACC pain scale [27].
4. Complications (major, minor), including any of the following within 30 days post- operation:

- Major: deep surgical site infection, wound dehiscence, anastomotic leak, bowel obstruction, pneumonia, sepsis, unplanned return to theatre, readmission to hospital within 30 days, mortality within 30 days
- Minor: superficial surgical site infection, perianal rash with skin breakdown, ileus, fever (above 38 degrees Celsius), vomiting, representation to hospital within 30 days
- Complications will be categorised as major or minor, and recorded as a yes/no event. If yes, the complications will be listed out.
5. Total analgesia requirement, including:

- Opioids (oral and IV, typically nurse-controlled analgesia). This will be recorded as a yes/no event and if yes, measured as the type, total mg/kg (as a central morphine unit [28]) and duration received post-operation,
- Oral simple analgesia (paracetamol, nurofen). This will be recorded as a yes/no event and if yes, measured as the type and number of doses.
6. Parent/guardian satisfaction, measured using the PedsQL Healthcare Satisfaction Generic Model [29].

### Participant timeline

Eligible participants will be recruited by an investigator and allocated after consent has been obtained from the parent/guardian (**Fig. 1**). Children allocated to the control group after their enterostomy closure will have a NGT remain in place post-operatively and will commence enteral nutrition after bowel function returns. Children in the intervention group will begin feeding within 24 hours after admission to the recovery room as tolerated. All participants will be closely monitored by the treating team and investigators for progress and complications. Patients will be discharged as per the treating team once on full enteral feeds, stooling, and pain-free or controlled on simple analgesia, with no parental or treating team concerns.

**Figure 1:**
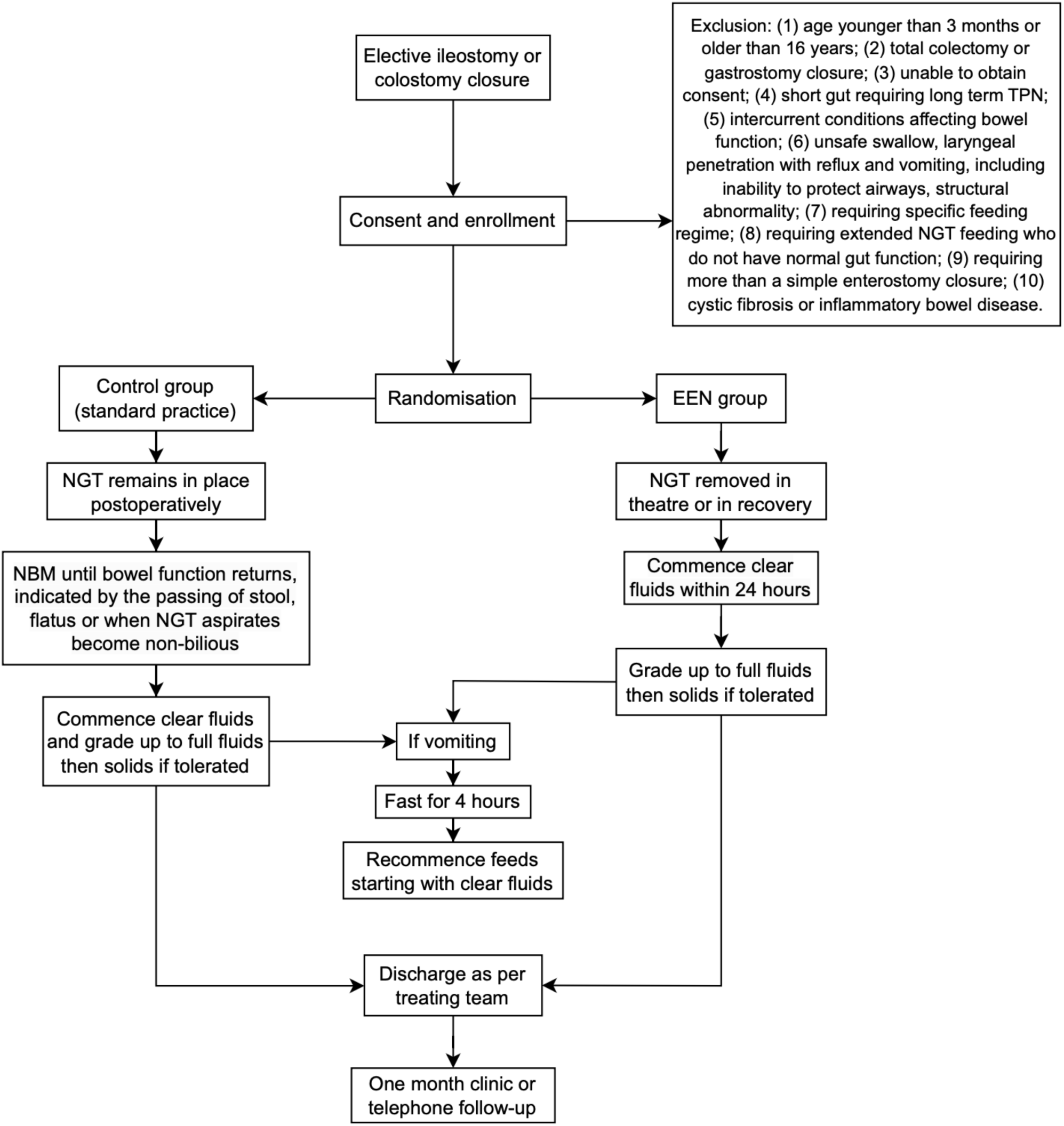
Study design and participant timeline. The figure outlines the study design and participant timeline from recruitment, randomisation, intervention to discharge. Abbreviations: **TPN**, total parenteral nutrition; **NGT**, nasogastric tube; **EEN**, early enteral nutrition; **NBM**, nil by mouth.

A parent/carer satisfaction survey will be performed at discharge (Online Supplementary Appendix 3). After one month, a telephone consult or clinic follow-up visit will be conducted by the treating team. Patients that experience any post-operative complications or concerns will be advised to present to the emergency department at either of the hospitals. Participants will be otherwise managed as per standard care procedures (**Table 1**).

**Table 1:**
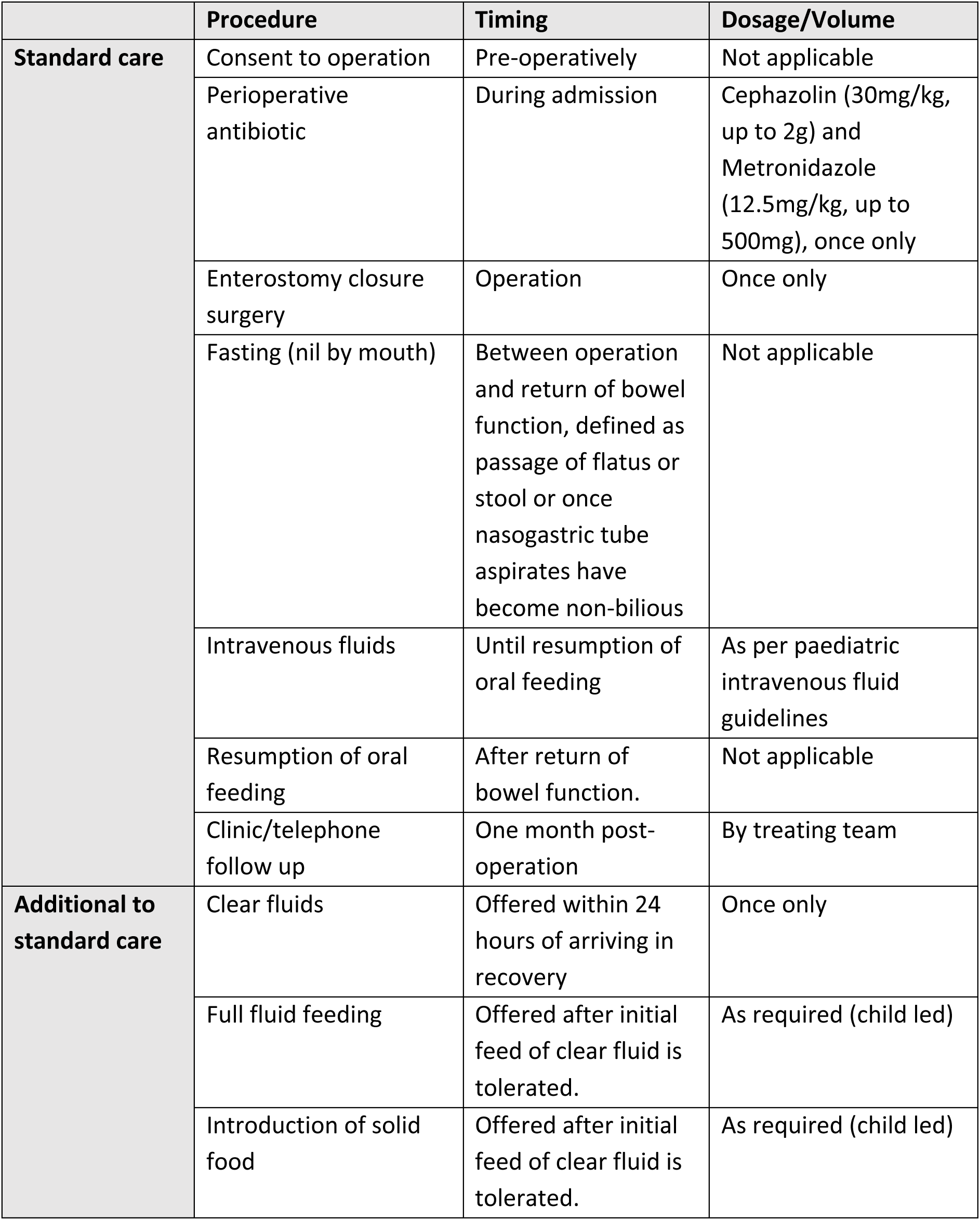
Standard care and additional to standard care procedures.

### Sample size and its justification

Sample size calculations were based on re-analysis of data from a previous retrospective cohort study conducted at the same study sites [26]. This study similarly examined the impact of EEN following stoma closure and reported the same primary and secondary outcomes. This data was re-analysed using restricted mean survival time (RMST). A power calculation was performed to obtain a minimally clinically important difference of discharge 24 hours earlier, at a time-point pre-specified at 168 hours (7 days), with power set at 80%. Based on this RMST analysis, we used a two-sample Z test to calculate a sample size of 68 patients (see Online Supplementary Appendix 5, section A for further justification). There are approximately 40 enterostomy closures across the two participating hospitals per year, of which we estimate up to 40% will be non-eligible due to the exclusion criteria, giving approximately 24 eligible cases per year. Assuming a recruitment rate of 50%, we would recruit 12 patients per annum, equating to 68 patients over five to six years.

### Data collection methods

Parents/carers will be given a study booklet to record the timing of feeds, passage of stool and vomiting during their hospital stay and a satisfaction form to complete at discharge (Online Supplementary Appendix 4). All other outcomes will be collected from the hospital’s electronic medical record. Outcome variables from this will be recorded in a password-protected Research Electronic Data Capture (REDCap) database [30]. Re-identifiable data will be uploaded by an investigator after the patient is discharged.

### Data management

Physical documents will be kept in a locked office and electronic data will be stored in REDCap. Deidentified data downloaded from REDCap for analysis will be stored on a password-protected computer secured behind a firewall.

### Statistical methods

Analysis of endpoint data will be performed based on the intention-to-treat principle, where participants are analysed in their randomised treatment group regardless of the treatment they received. For continuous variables, characterisation will be by mean and standard deviation for normally distributed data or median and interquartile range for non-normally distributed data. Categorical variables will be described using frequencies and percentages.

Time-based variables including the primary outcome of LOS, time to feeds (clear fluids, free fluids, full feeds) and time to passage of stool will be presented using Kaplan-Meier curves and analysed using RMST. Sensitivity analyses will include Cox proportional hazards models and unadjusted models, reported with 95% confidence interval and p-values (see Online Supplementary Appendix 5, section B for details of primary analysis).

Other secondary outcomes (pain score, complications, total analgesia, parent satisfaction) will be analysed using appropriate regression models based on the type and distribution of each variable. This may include logistic regression for binary outcomes, Poisson or negative binomial models for counts, and Gaussian models for continuous variables. Where required, robust methods like bootstrapping or robust variance estimators will be applied (see Online Supplementary Appendix 5, section C for model diagnostics and selection).

Secondary outcomes will be considered exploratory. Accordingly, p-values will be reported to aid interpretation rather than for formal hypothesis testing. To minimise the risk of type II errors, no adjustments will be made for multiple comparisons, particularly for serious complications, where failing to detect a true difference could have important clinical implications and may be more consequential than a false positive. Model selection techniques will not be used to determine which covariates to adjust for. Any adjusted analyses will be prespecified based on clinical relevance and reported as sensitivity analyses to explore robustness of the findings and inform future studies.

### Withdrawal of consent and missing data

The total number of consent withdrawals after randomisation will be reported but their data not included in the analysis.

All data are recorded in the medical records aside from the parent/care satisfaction, time to feeds, stools and vomits, and the satisfaction survey which is completed prior to discharge. Therefore, we anticipate minimal missing data for the primary endpoint and secondary outcomes. However, if missing data does occur, we will analyse the data based on our assessment of the patterns and predictors of missingness.

Missing baseline covariate data will be assessed based on *a priori* theoretical considerations regarding the data generating and collection processes. This will be supported by exploratory analyses, including logistic regression models to identify potential predictors of missingness.

The primary analysis will assume that missing data are Missing at Random (MAR) and will be handled using Multiple Imputation (MI), rather than complete case analysis, to increase statistical power. Imputation will include all relevant covariates, treatment group, hospital and outcome variables. Analyses will be conducted using standard procedures, and results combined using Rubin’s rules (see Online Supplementary Appendix 5, section D for MI process and software).

For the time-based variables (LOS, time to clear fluids, free fluids, full feeds, passage of stool), censored observations will be incorporated into the analyses under the assumption of Censoring at Random. Where there are concerns about informative censoring, we will perform sensitivity analyses which may include a competing risk analysis like the Fine-Gray model, if appropriate (see Online Supplementary Appendix 5, section E for handling and analysis of censored data).

Secondary outcomes (post-operative pain score, complications, total analgesia, parent satisfaction) will also be analysed under the MAR assumption.

There is the possibility that some data may be Missing Not at Random (MNAR). Since a formal MNAR analysis is not planned for this study, any potential bias from MAR violations will be addressed as a limitation in the discussion of the final study report.

## MONITORING

### Data monitoring

As the study is not examining major health outcomes and the intervention is low risk, a stand- alone data monitoring and safety committee is not required [31]. However, as recommended by the National Health and Medical Research Council (NHMRC), a committee consisting of an independent dietician, surgeon and statistician has been established to meet annually and provide oversight, review recruitment, safety, data quality, protocol adherence and retention [31].

### Harms and adverse event reporting

Most of the patient care is usual clinical care, including the preoperative care, surgical approach, post-operative care (aside from feeding) and medications. Participants will be closely monitored throughout their hospital stay to ensure early detection and management regarding possible complications that can be associated with early feeding or stoma closure, noting the literature reports that most complications are of no higher prevalence than with standard care [32]. The one exception is a higher chance of vomiting with early feeding [11]. This risk will be included in the parent information provided prior to enrolment into the study. For orally fed children, feeding will be guided by the child’s own desire for oral intake. In this context, vomiting may produce some discomfort but is very unlikely to represent a material risk to the conscious child who meets inclusion criteria. For children who otherwise meet inclusion criteria but are fed through NGT (e.g. oral aversion), initial feeds will be given while awake, starting with clear fluids within 24 hours and progressing to full fluids within 48 hours, according to the timing in the study protocol, with observation for tolerance. The risk associated with vomiting is further mitigated through the exclusion of children with swallowing abnormalities or problems with protecting their airway during feeding or vomiting. If vomiting occurs, the child will be assessed by a member of the treating team to ensure that continuing feeds is not a risk. This involves excluding bowel obstruction, ileus or systemic symptoms precluding safe feeding, such as decreased level of consciousness. All serious adverse effects reported by the subject or observed by the investigators or hospital staff will be documented and reported to the SCHN Ethics Committee and according to the NSW Health “Incident Management” Policy Directive [33].

### Auditing

The study team will meet fortnightly to review recruitment, data completion and any withdrawals and protocol violations. Data will be audited by the research team and reviewed by the statistician every six to 12 months, depending on recruitment rate.

## ETHICS AND DISSEMINATION

This protocol has been approved by The Sydney Children’s Hospital Network Human Research Ethics Committee (SCHN HREC; 2021/ETH01062).

### Protocol amendments

Permission for amendments to the protocol will be sought from the SCHN HREC. Any difference that may impact the participants will be relayed to them and consent renewal will be obtained appropriately. Any further publications will include updated trial registries and protocol amendments.

### Confidentiality

Data will be deidentified by establishing a master document in REDCap that will have the patient’s name, medical record number and study number recorded. A separate main study database will use the study number. Any download files will be deidentified and stored in password-protected firewalled servers.

As per the NHMRC [34], data will be securely stored for 15 years after the study concludes or until the youngest participant reaches the age of 25 (whichever comes later). Following this, electronic data will be erased and physical copies shredded securely.

No tissue collection or bio-banking outside the typical clinical care will occur in this study.

### Access to data

The primary investigator will have access to the final trial dataset.

### Ancillary data

There will be no ancillary data collected from the participants.

### Dissemination policy

The trial is registered on ANZCTR (ACTRN12621001414808), which has open access. To ensure that the findings are distributed to relevant healthcare practitioners, it will be published in an appropriate peer-reviewed journal and may be presented in local conferences or meetings.

The investigators will distribute the final report to the participants who wish to know the results.

## EXPECTED OUTCOMES AND SIGNIFICANCE OF THE RESEARCH PROJECT

There are currently no high-quality studies assessing the benefits of EEN in paediatric patients undergoing general surgical procedures. Existing studies are retrospective or include a wide range of disparate conditions and feeding regimes. This project will be the first well-designed Australian RCT to compare EEN with standard clinical practice after enterostomy closure in the paediatric setting. It will also be the first to include consumer outcomes and an assessment of the effect of EEN on post-operative pain. The findings will provide important information for the application of EEN and broader post-operative recovery protocols in paediatric clinical practice.

## Data Availability

All data produced in the present study are available upon reasonable request to the authors. This protocol paper illustrates an ongoing RCT so the data we have obtained is incomplete.

## AUTHORS’ CONTRIBUTIONS

### Contributors

Joyce Ly: project administration, writing (original draft).

James Cope: conceptualisation, methodology, project administration, writing (review and editing).

Nicholas Olsen: methodology (sample size, statistical methods, missing values), writing (review and editing).

Soundappan SV Soundappan: supervision, methodology, project administration, resources. Susan Adams (guarantor): supervision, conceptualisation, methodology, project administration, resources, writing (review and editing).

## REFERENCES

1. Behera BK, Misra S, Tripathy BB. Systematic review and meta-analysis of safety and efficacy of early enteral nutrition as an isolated component of Enhanced Recovery After Surgery [ERAS] in children after bowel anastomosis surgery. Journal of Pediatric Surgery. 2022;57(8):1473–9.

2. Amanollahi O, Azizi B. The comparative study of the outcomes of early and late oral feeding in intestinal anastomosis surgeries in children. African Journal of Paediatric Surgery. 2013;10(2):74–7.

3. Ekingen G, Ceran C, Guvenc BH, et al. Early enteral feeding in newborn surgical patients. Nutrition. 2005;21(2):142–6.

4. Shang Q, Geng Q, Zhang X, et al. The impact of early enteral nutrition on pediatric patients undergoing gastrointestinal anastomosis a propensity score matching analysis. Medicine (United States). 2018;97(9).

5. Sunstrom R, Hamilton N, Fialkowski E, et al. Minimizing variance in pediatric gastrostomy: does standardized perioperative feeding plan decrease cost and improve outcomes? The American Journal of Surgery. 2016;211(5):948–53.

6. Yadav PS, Choudhury SR, Grover JK, et al. Early feeding in pediatric patients following stoma closure in a resource limited environment. Journal of Pediatric Surgery. 2013;48(5):977–82.

7. Ross AR, Hall NJ, Ahmed SA, et al. The extramucosal interrupted end-to-end intestinal anastomosis in infants and children; a single surgeon 21year experience. Journal of Pediatric Surgery. 2016;51(7):1131–4.

8. McClave SA, Heyland DK. The physiologic response and associated clinical benefits from provision of early enteral nutrition. Nutrition in Clinical Practice. 2009;24(3):305–15.

9. Kehlet H. Multimodal approach to control postoperative pathophysiology and rehabilitation. British Journal of Anaesthesia. 1997;78(5):606–17.

10. Herbert G, Perry R, Andersen HK, et al. Early enteral nutrition within 24 hours of lower gastrointestinal surgery versus later commencement for length of hospital stay and postoperative complications. Cochrane Database of Systematic Reviews. 2019;2019(7).

11. Lewis SJ, Egger M, Sylvester PA, et al. Early enteral feeding versus “nil by mouth” after gastrointestinal surgery: systematic review and meta-analysis of controlled trials. BMJ. 2001;323(7316):773.

12. Kiyama T, Onda M, Tokunaga A, et al. Effect of early postoperative feeding on the healing of colonic anastomoses in the presence of intra-abdominal sepsis in rats. Diseases of the Colon & Rectum. 2000;43(10):S54–S8.

13. Tadano S, Terashima H, Fukuzawa J, et al. Early Postoperative Oral Intake Accelerates Upper Gastrointestinal Anastomotic Healing in the Rat Model. Journal of Surgical Research. 2011;169(2):202–8.

14. Udén P, Blomquist P, Jiborn H, et al. Impact of long-term relative bowel rest on conditions for colonic surgery. The American Journal of Surgery. 1988;156(5):381–5.

15. Moss G, Greenstein A, Levy S, et al. Maintenance of GI Function after Bowel Surgery and Immediate Enteral Full Nutrition. I. Doubling of Canine Colorectal Anastomotic Bursting Pressure and Intestinal Wound Mature Collagen Content. Journal of Parenteral and Enteral Nutrition. 1980;4(6):535–8.

16. Davila-Perez R, Bracho-Blanchet E, Galindo-Rocha F, et al. Early feeding vs 5-day fasting after distal elective bowel anastomoses in children. A Randomized Controlled Trial. Surgical Science. 2013;4:45–8.

17. Ghosh A, Biswas SK, Basu KS, et al. Early Feeding after Colorectal Surgery in Children: Is it Safe? Journal of Indian Association of Pediatric Surgeons. 2020;25(5):291–6.

18. Iqbal MJ, Iqbal A, Anwer MS, et al. Early versus delayed feeding in pediatric patients following stoma reversal in a resource limited environment. Iranian Journal of Pediatric Surgery Vol. 2020;6(2):59–65.

19. Peng Y, Xiao D, Xiao S, et al. Early enteral feeding versus traditional feeding in neonatal congenital gastrointestinal malformation undergoing intestinal anastomosis: A randomized multicenter controlled trial of an enhanced recovery after surgery (ERAS) component. Journal of Pediatric Surgery. 2021;56(9):1479–84.

20. Santos-Jasso KA, Lezama-Del Valle P, Arredondo-Garcia JL, et al. Efficacy and safety of an abbreviated perioperative care bundle versus standard perioperative care in children undergoing elective bowel anastomoses: A randomized, noninferiority trial. Journal of Pediatric Surgery. 2020;55(10):2042–7.

21. Paul SK, Biswas I, Howlader S, et al. Early enteral feeding versus traditional feeding after colostomy closure in paediatric patients: A comparative study of postoperative outcome. Faridpur Medical College Journal. 2015;10(1):29–32.

22. Prasad GR, Rao JS, Aziz A, et al. Early enteral nutrition in neonates following abdominal surgery. Journal of Neonatal Surgery. 2018;7(2):21.

23. Ashjaei B, Ghamari Khameneh A, Darban Hosseini Amirkhiz G, et al. Early oral feeding versus traditional feeding after transanal endorectal pull-through procedure in Hirschsprung’s disease. Medicine (Baltimore). 2019;98(10):e14829.

24. Gil-Vargas M, Saavedra-Pacheco M, Coral-García M. Early feeding versus traditional feeding in children with ileostomy closure. Journal of Indian Association of Pediatric Surgeons. 2022;27(2):223–6.

25. Sangkhathat S, Patrapinyokul S, Tadyathikom K. Early enteral feeding after closure of colostomy in pediatric patients. Journal of Pediatriac Surgery. 2003;38(10):1516–9.

26. Cope J, Greer D, Soundappan SSV, et al. The Safety and Efficacy of Early Enteral Nutrition After Paediatric Enterostomy Closure - The EPOC Study. Journal of Pediatric Surgery. 2024;59(4):701–8.

27. Merkel SI, Voepel-Lewis T, Shayevitz JR, et al. The FLACC: a behavioral scale for scoring postoperative pain in young children. Journal of Pediatric Nursing. 1997;23(3):293–7.

28. Adams MC, Sward KA, Perkins ML, Hurley RW. Standardizing research methods for opioid dose comparison: the NIH HEAL morphine milligram equivalent calculator. Pain. 2025;166(8):1729–37.

29. Varni JW, Seid M, Rode CA. The PedsQL™: Measurement Model for the Pediatric Quality of Life Inventory. Medical Care. 1999;37(2).

30. Harris PA, Taylor R, Thielke R, et al. Research electronic data capture (REDCap)—A metadata-driven methodology and workflow process for providing translational research informatics support. Journal of Biomedical Informatics. 2009;42(2):377–81.

31. National Health and Medical Research Council. Data safety monitoring boards (DSMBs) [online]. 2018. https://www.nhmrc.gov.au/sites/default/files/documents/reports/data-safety-monitoring-boards.pdf (accessed 9 April 2025).

32. Greer D, Karunaratne YG, Karpelowsky J, et al. Early enteral feeding after pediatric abdominal surgery: A systematic review of the literature. Journal of Pediatric Surgery. 2020;55(7):1180–7.

33. New South Wales Health. Incident management [online]. 2020. https://www1.health.nsw.gov.au/pds/ActivePDSDocuments/PD2020_047.pdf (accessed 27 January 2025).

34. National Health and Medical Research Council (NHMRC). National statement on ethical conduct in human research. Canberra (AU): National Health and Medical Research Council; 2023.

