## Supplementary material for "Early enteral nutrition after Paediatric Ostomy Closure (EPOC): a protocol for a multicentre, prospective randomised controlled trial": Online Supplementary Appendix 1

**Information Sheet**

| | |
|---|---|
| **Study Title** | Early enteral nutrition after Paediatric Ostomy Closure (EPOC): A Prospective Randomised Controlled Trial |
| **Principal Investigator/s** | Susan Adams - Department of Paediatric Surgery, Sydney Children's Hospital, Randwick. 9382 1776<br>Soundappan Soundappan - Department of Paediatric Surgery, Children's Hospital at Westmead. 9845 3235 |
| **Main Study Contact Person** | Susan Adams - 9382 1776 |

### 1. Introduction

Your child is invited to take part in a research study titled *Early enteral nutrition after Paediatric Ostomy Closure (EPOC): A Prospective Randomised Controlled Trial***.** This study will be conducted within the Department of Paediatric Surgery, Sydney Children's Hospital Network, Randwick and Westmead campuses.

Your child is invited because they are booked into hospital to have their stoma (ostomy) closed.

This information sheet tells you about the study. It explains the processes involved with taking part. Knowing what is involved will help you decide if you and your child want to take part in the study. Please read this information carefully. Ask questions about anything that you don't understand or want to know more about.

Participation in this research is voluntary. If you or your child do not wish to take part, you do not have to. It will not affect your care at the hospital whether or not you decide to take part.

### 2. What is the purpose of this study?

Following the closure of a stoma, post-operative fasting for a few days has been the norm. It is common for the child to remain fasted (not have anything to eat or drink) until they have passed wind or opened their bowels, which may take some days. This is called 'usual practice'. However, this prolonged fasting may not be necessary. We now know from research in adults and some reports in children that early feeding after abdominal surgery is associated with shorter length of stay in hospital and is not associated with higher risks of complications or other adverse outcomes.

This study aims to assess if early feeding after an elective stoma closure procedure will decrease pain medication (analgesia) requirements, complications, length of stay and increase patient satisfaction. This will be done by comparing children in two groups

1. Early Feeding: those who have their stoma closed and commence feeding within 24 hours following surgery.

2. Usual Practice - Feeding after Fasting: those who have their stoma closed then fast until they pass wind or open their bowels and/or until the surgeon is happy their tummy is back to normal. This is what is usually done.

Participants will be assigned randomly to be treated with either early feeding or feeding after a fasting period. You cannot choose which group your child will be in.

### 3. Why have I been invited to this study?

Your child is invited to take part in this study because they are booked to have an elective stoma closure operation at Sydney Children's Hospital Randwick/Westmead Children's Hospital.

### 4. Do I have to take part in this study?

Participation in any research project is voluntary. If you or your child do not wish to take part, you do not have to. If you decide to take part and later change your mind, you are free to withdraw from the project at any stage.

Your decision whether to take part or not to take part, or to take part and then withdraw, will not affect your child's routine care, your relationship with professional staff or your relationship with Sydney Children's Hospital Randwick/Westmead Children's Hospital. If you do decide to take part, you will be given a consent form to sign and you will be given a copy to keep.

### 5. What does participation in this study involve?

When your child is booked to come into hospital to have their stoma closed, your doctor will decide if your child is suitable for inclusion this study. At that point you and your child will be invited to take part in this study.

If you decide to take part, your child will be allocated randomly to one of two groups of the study.

**GROUP 1**: Early Feeding: Oral feeds will be offered within 24 hours of the operation.

**GROUP 2:** Usual practice: Feeding after Fasting - feeds commenced after the doctors assess that your child's tummy is getting back to normal.

If your child is in Group 1 - Early Feeding - they will be offered clear fluids or breast milk as their first feed within 24 hours of the operation. After that, they can drink other fluids (such as formula, milk, yoghurt, depending on the usual diet they were on before the operation). Solid food (for babies and children who usually eat solid food) will then be offered to your child within 48 hours of the operation.

If your child is in Group 2 – Usual Practice - Feeding after Fasting - they will be offered feeds after the surgeon assesses them and decides that their tummy is getting back to normal, such as after they have passed wind or stool. This varies from child to child but may take a couple of days.

For both groups, if vomiting occurs, your child will fast for 4 hours and then, as long as they are OK, start with some clear fluids again and then be offered other fluid and food as above.

Aside from the feeding, both groups will be cared for the same way. All children will be closely monitored. At the time of discharge from hospital, we will ask you to fill out a parent/carer satisfaction survey. Following discharge, we will organise a telephone interview or clinical follow up visit for 1 month post discharge.

As is usual advice when going home from hospital, children that experience any post-operative complications or concerns will be advised to present to the emergency department and Sydney Children's Hospital/Children's Hospital at Westmead or their local emergency department if they live along way away.

Information collected about your child during the study period includes;

- Age
- Gender
- Location of enterostomy (ileostomy, colostomy)
- Hours to first feed (a) clear fluids (b) full fluids (c) solids (d) eating as normal
- Time to pass stool
- Pain relief used
- Pain score - to assess how much pain after the operation
- Temperature
- Length of stay
- Information about any complications
- Whether you child came back to the emergency department
- Whether you child was readmitted after having been discharged
- Whether they required further operations or procedures
- Carer/parent satisfaction survey at discharge from hospital

**A table explaining the study:**

| | |
|---|---|
| **Step 1: Eligibility** | We will assess if your child is eligible to be included in the study |
| **Step 1: Recruitment** | We will ask if you would like to take part. We will answer any questions you may have and run you through what the study will involve. |
| **Step 2: Allocation** | We will randomly allocate your child to either Group 1 or Group 2. |
| **Step 3: Treatment** | Depending on which group your child is in, they will either receive Feeding after Fasting (after return of bowel function) or they will receive Early Feeding - within 24 hours after your surgery. |
| **Step 4: Follow up** | We will organise for a telephone interview or clinical follow up visit 1 month after your child is discharged from hospital. |

### 6. What are the possible risks and disadvantages of taking part?

The risks of this study are very low. Most of the patient care is standard care, including the pre-operative care, the operation, the post-operative care (aside from feeding) and medications, including pain medications. Children in both groups will be closely monitored throughout hospital stay to make sure everything is going well and to detect any problems that may need addressing.

Complications can rarely be associated with stoma closure, regardless of the timing of feeding. Vomiting can also happen after an operation, regardless of the timing of feeds. It is possible that those children that are fed early, may have a higher change of vomiting. If your child vomits, they will be assessed by a member of the surgical team to make sure they are OK. As long as they are, they'll have a rest for a while, then start feeds again once the vomiting has settled. Any problems detected will be looked after as we usually do.

### 7. What are the possible benefits of taking part?

There are no specific benefits to your child in taking part in this study. We hope that the results from this study will help find out if children who come into hospital to have their stoma closed can benefit from feeding early.

### 8. What will happen to my child's information?

By signing the consent form, you consent to the research team collecting and using personal information about your child for the research project. Their privacy and confidentiality will be protected at all times. Your child's information will only be used for the purpose of this research study and it will only be disclosed with your permission, except as required by law. For example, researchers are required to report if a participant is believed to be at risk of harm.

The information collected will be kept on a secure password protected on-line database at the University of New South Wales that is designed especially for this sort of research. In order to protect your child's privacy, the study team will remove any information that may be used to identify them from any study documents, and instead of their name appearing on the documents, they will be identified by a specific study code number that applies only to them. Only this code number will be used on any research-related information collected about them for this study, so that their identity as part of the study will be kept completely private. Only the study team at Sydney Children's Hospital Randwick and Westmead Children's Hospital will have the ability to link this code number with their personal information. Your child's data will be stored for 15 years after the study finishes, or until the youngest child in the study turns 25, whichever of the two is longer.

We also seek your consent to use results of this study to contribute to further work in the area of Enhanced Recovery After Surgery (ERAS). Research in this field is designed at improving post-operative recovery without increasing complications, ultimately aimed at improving overall patient care. Any information regarding you or your child would be de-identified and be kept completely confidential. Any further research conducted would also only be done with approval from the Sydney Children's Hospital Network ethics board.

If you and your child withdraw from the study, we will not collect any more information about them. We would like to keep the information we have already collected about them to help us ensure that the results of the research project can be measured properly. Please let us know if you do not want us to do this.

### 9. How will the results of the study be distributed?

It is anticipated that the results of this research project will be published in a medical journal and/or presented at scientific and educational meetings. In any publication and/or presentation, information will be presented in such a way that your child cannot be identified, except with your expressed permission.

You can indicate on the consent form if you wish to receive a summary of the study findings.

### 10. Who should I contact if I have any questions?

If you have any questions or want more information about this study before or during participation, you can contact:

| | |
|---|---|
| Susan Adams | Department of Paediatric Surgery, Sydney Children's Hospital, Randwick, 9382 1776 |
| Soundappan Soundappan | Department of Paediatric Surgery, Children's Hospital at Westmead, 9845 3235 |

### 11. Who do I contact if I have concerns about the study?

All research in Australia involving humans is reviewed by an independent group of people called a Human Research Ethics Committee (HREC). This study has been approved by the Sydney Children's Hospitals Network (SCHN) HREC **(approval number: ETH01062)**.

If you have any concerns or complaints about any aspect of the project or the way it is being conducted, you may contact the Executive Officer of the SCHN HREC on (02) 9845 1253 or.

*This Information Sheet is for you to keep. We will also give you a copy of the signed consent form.*

### Parent / Guardian Consent Form

| Study Title | Early enteral nutrition after Paediatric Ostomy Closure (EPOC): A Prospective Randomised Controlled Trial |
|---|---|
| Principal Investigator/s | Susan Adams - Department of Paediatric Surgery, Sydney Children's Hospital, Randwick. 9382 1776<br>Soundappan Soundappan - Department of Paediatric Surgery, Children's Hospital at Westmead, 9845 3235 |
| Main Study Contact Person | Susan Adams - 9382 1776 |

*Declaration by Parent / Guardian*

☐ I have read the Parent / Guardian Information Sheet or someone has read it to me in a language that I understand.

☐ I understand the purposes, procedures and risks of the research project described in the Parent / Guardian Information Sheet.

☐ I have had an opportunity to ask questions and I am satisfied with the answers I have received.

☐ I freely agree to the child participating in this research project as described and understand that I am free to withdraw them at any time during the project without affecting their future health care.

☐ I understand that I will be given a signed copy of this document to keep.

☐ I give permission for my child's treating doctor, other health professionals, hospitals or laboratories outside this hospital to release information to UNSW concerning my child's condition and treatment for the purposes of this project. I understand that such information will remain private and confidential.

☐ I understand that the results of this project, may be given for the use of data in future research projects that are: an extension of the original project; or in the same general area of research.

☐ I wish to receive a lay summary of the study findings via the following email / post address:

______________________________________________________________

Name of Child (please print): ______________________________________________________

Signature of Child: ________________________________ Date: ____________________

Name of Parent / Guardian (please print): _____________________________________________

Signature of Parent / Guardian: _________________________ Date: ____________________

*Under certain circumstances (see Note for Guidance on Good Clinical Practice CPMP/ICH/135/95 at 4.8.9) a witness* to informed consent is required.*

Name of Witness* to Parent / Guardian's Signature (please print): ______________________________

Signature of Witness: ___________________________________ Date: _____________________

* The Witness is not to be the investigator, a member of the study team or their delegate. In the event that an interpreter is used, the interpreter may not act as a witness to the consent process. Witnesses must be over 18 years of age
