## Supplementary material for "Early enteral nutrition after Paediatric Ostomy Closure (EPOC): a protocol for a multicentre, prospective randomised controlled trial": Online Supplementary Appendix 3

### Parent satisfaction form

Please answer the following questions telling us how happy you are with the care you, your child, and your family have received at the hospital and after discharge from hospital from the staff. Please tell us how happy you are with each item by circling:

**0** if you are **never** happy

**1** if you are **sometimes** happy

**2** if you are **often** happy

**3** if you are **almost always** happy **4** if you are **always** happy

There are no right or wrong answers. Please circle **N/A** (**not applicable**) if the item does not apply to you.
If you do not understand a question, please ask for help by contacting the investigators on the information sheet you have received

#### How happy are you with…?

| **INFORMATION** | **Very unhappy** | **Somewhat unhappy** | **Somewhat happy** | **Happy** | **Very happy** | **Not Applic** |
|---|---|---|---|---|---|---|
| 1. How much information was provided to you about your child's diagnosis | 0 | 1 | 2 | 3 | 4 | N/A |
| 2. How much information was provided to you about the options for treatment of your child's health condition | 0 | 1 | 2 | 3 | 4 | N/A |
| 3. How much information was provided to you about the possible side effects of your child's treatment | 0 | 1 | 2 | 3 | 4 | N/A |
| 4. How soon information was given to you about your child's test results | 0 | 1 | 2 | 3 | 4 | N/A |
| 5. How often you are updated about your child's health | 0 | 1 | 2 | 3 | 4 | N/A |

the children's hospital at Westmead + SYDNEY CHILDREN'S HOSPITAL RANDWICK = The Sydney Children's Hospitals Network (Randwick and Westmead)

#### How happy are you with…?

| **Inclusion of Family** | Very unhappy | Somewhat unhappy | Somewhat happy | Happy | Very happy | Not Applic |
|---|---|---|---|---|---|---|
| 1. The sensitivity shown to you and your family during your child's treatment | 0 | 1 | 2 | 3 | 4 | N/A |
| 2. The willingness to answer questions that you and your family may have | 0 | 1 | 2 | 3 | 4 | N/A |
| 3. The effort to include your family in discussion of your child's care and other information about your child's health condition | 0 | 1 | 2 | 3 | 4 | N/A |
| 4. How much time the staff gave you to ask any questions you may have had about your child's health condition and treatment | 0 | 1 | 2 | 3 | 4 | N/A |

#### How happy are you with…?

| **Communication** | Very unhappy | Somewhat unhappy | Somewhat happy | Happy | Very happy | Not Applic |
|---|---|---|---|---|---|---|
| 1. How well the staff explained your child's health condition and treatment to **your child** in a way that she/he could understand | 0 | 1 | 2 | 3 | 4 | N/A |
| 2. The time taken to explain your child's health condition and treatment to **you** in a way you could understand | 0 | 1 | 2 | 3 | 4 | N/A |
| 3. How well the staff listens to you and your concerns | 0 | 1 | 2 | 3 | 4 | N/A |
| 4. The preparation provided for **you** about what to expect during the trial | 0 | 1 | 2 | 3 | 4 | N/A |
| 5. The preparation provided for **your child** about what to expect during the trial | 0 | 1 | 2 | 3 | 4 | N/A |

How happy are you with…?

| Technical Skills | Very unhappy | Somewhat unhappy | Somewhat happy | Happy | Very happy | Not Applic |
|---|---|---|---|---|---|---|
| 1. How well the staff responds to your child's needs | 0 | 1 | 2 | 3 | 4 | N/A |
| 2. Efforts to keep your child comfortable and as pain-free as possible | 0 | 1 | 2 | 3 | 4 | N/A |
| 3. How much time the staff took to help you with your child coming back home | 0 | 1 | 2 | 3 | 4 | N/A |

How happy are you with…?

| Emotional Needs | Very unhappy | Somewhat unhappy | Somewhat happy | Happy | Very happy | Not Applic |
|---|---|---|---|---|---|---|
| 1. The amount of time given to your child to play, talk about her/his feelings, and any questions she/he may have | 0 | 1 | 2 | 3 | 4 | N/A |
| 3. The amount of time spent helping your child with going back to day care/preschool/school | 0 | 1 | 2 | 3 | 4 | N/A |
| 3. The amount of time spent attending to **your child's** emotional needs | 0 | 1 | 2 | 3 | 4 | N/A |
| 4. The amount of time spent attending to **your** emotional needs | 0 | 1 | 2 | 3 | 4 | N/A |

How happy are you with…?

| Overall Satisfaction | Very unhappy | Somewhat unhappy | Somewhat happy | Happy | Very happy | Not Applic |
|---|---|---|---|---|---|---|
| 1. The overall care your child received during the trial | 0 | 1 | 2 | 3 | 4 | N/A |
| 2. How friendly and helpful the staff were | 0 | 1 | 2 | 3 | 4 | N/A |
| 3. The way your child was treated at the hospital | 0 | 1 | 2 | 3 | 4 | N/A |

Please tell us to what degree you agree with the following statements by circling

**0** if you strongly disagree

**1** if you disagree

**2** if you are **neutral – neither agree nor disagree**

**3** if you agree

**4** if you strongly agree

Please circle **N/A** (**not applicable**) if the item does not apply to you.
If you do not understand a question, please ask for help by contacting the investigators on the information sheet you have received

### *Do you agree with the statements…?*

| TRIAL OUTCOMES | Strongly Disagree | Disagree | Neutral | Agree | Strongly Agree | Not Applic |
|---|---|---|---|---|---|---|
| 1. The information provided to me at the start about the trial prepared me for participation. | 0 | 1 | 2 | 3 | 4 | N/A |
| 2. At the start of the trial, I felt anxious about the outcome for my child | 0 | 1 | 2 | 3 | 4 | N/A |
| 3. I am satisfied with my child's recovery | 0 | 1 | 2 | 3 | 4 | N/A |
| 4. I am satisfied with the length of time my child was required to fast after the operation (not have anything to drink or eat) | 0 | 1 | 2 | 3 | 4 | N/A |
| 5. I am confident that the right decisions for my child were made during the study | 0 | 1 | 2 | 3 | 4 | N/A |
| 6. I worry that my child will need further surgery/treatment in the future | 0 | 1 | 2 | 3 | 4 | N/A |
| 7. All of my concerns during the study were addressed | 0 | 1 | 2 | 3 | 4 | N/A |

***Please add any further comments about your child's care during the trial and their hospital stay***

Thank you for your participation!
