## Supplementary material for "Early enteral nutrition after Paediatric Ostomy Closure (EPOC): a protocol for a multicentre, prospective randomised controlled trial": Online Supplementary Appendix 4

### Online Supplementary Appendix 4: Sample size, statistical methods and missing data

#### Section A: Sample size justification

Sample size calculations were based on re-analysis of data from a previous retrospective cohort study conducted at the same study sites (Cope et al., 2024). This study similarly examined the impact of EEN following stoma closure and reported the same primary and secondary outcomes. This data was re-analysed using restricted mean survival time. A power calculation was performed to obtain a minimally clinically important difference (MCID) of discharge 24 hours earlier, at a time-point (tau) pre-specified at 168 hours (7 days) with power set at 80%. Based on this restricted mean survival time (RMST) analysis, we used a two-sample Z-test to calculate a sample size of 68 (34 per group). We believe that this is both an important and realistic MCID target. Our analysis found that the RMST difference between groups at 7 days was discharge 25.33 hours earlier (SE 8.506, 95% CI -42.003 to -8.658 hours, p = 0.003). This result was obtained after adjustment for age and stoma location – noting that as this retrospective data was from one site, hospital was not included as a covariate. The timepoint of 7 days was chosen as most patients would be discharged by this timepoint. We chose RMST as the analysis strategy over Cox regression, as a hazards ratio tells us nothing about the difference in time between groups, only the ranking of events.

The retrospective study used for sample size calculation can be found at: https://www.jpedsurg.org/article/S0022-3468(23)00695-4/fulltext.

#### Section B: Primary analysis

Time-based variables including the primary outcome of LOS, time to feeds (clear fluids, free fluids, full feeds) and time to passage of stool will be presented using Kaplan-Meier curves and analysed using RMST, with tau defined at 168 hours (7 days), assuming at least one participant remains at risk in each group. If this condition is not met, tau will be the maximum timepoint at which at least one participant remains in the risk set in each group. Analyses will be conducted using the *survRM2* package in R. The primary outcome will be the adjusted difference in RMST between groups, reported with 95% confidence interval (CI) and p-value. We will also report the RMST ratio (treatment vs. control) and the difference in restricted mean time lost (RMTL), with 95% CI and p-value. These will be considered supportive to aid interpretation.

Sensitivity analysis will include an unadjusted RMST model (treatment only) with group-specific survival times and differences reported with 95% CI. This serves two purposes: (i) RMST estimates per group cannot be obtained with covariate adjustments in *survRM2*, and (ii) to assess sensitivity to model specification. Given that our prior retrospective study found no effect of age (p = 0.59) or stoma location (p = 0.98) on LOS, it is plausible that that these covariates do not influence the outcome. If this is true, the treatment effect in the primary model remains unbiased. However, the inclusion of unnecessary covariates would reduce statistical power and precision of the estimated treatment effects.

Further sensitivity analyses will use Cox proportional hazards models with the same covariates as the primary model, stratified by hospital, to provide hazard ratios with 95% CI alongside RMST results. We will also report a Cox model with treatment as the sole predictor, stratified by hospital. The proportional hazards assumption will be formally assessed with Schoenfeld residual plots and hypothesis testing using the *cox.zph* command in R. If any covariates violate the proportional hazards assumption, they will be incorporated as stratification variables using the *strata* option, allowing for separate baseline hazard functions for each level of that covariate. In this scenario, the RMST difference (95% CI, p-value) will be reported as the primary measure, with the RMST ratio and RMTL difference provided as supportive outcomes.

**Section C: Model diagnostics and selection**

For other secondary outcomes, analysis will be performed with generalised linear models, which may include beta regression for bounded scales, Poisson or negative-binomial regression for count data, logistic regression for binary outcomes, or Gaussian models where appropriate. Prior to viewing model results, the distributions and link functions for secondary outcomes will be obtained empirically by a statistician, based on inspection of residual versus predicted plots, QQ residual plots, other common model diagnostic tests, and theoretical considerations of the underlying data generating process. If model assumptions are not met with generalised linear models, options such as bootstrapping or robust (sandwich) variance estimators will be used for violations of heteroscedasticity. Given the expected sample size, minor to moderate violations of normality of residuals is acceptable due to the central limit theorem. For severe departures from normality or poor residual fits, transformation of the response variables or use of different distributions and link functions may also be employed. In cases of binary outcomes with perfect prediction, both Firth's penalised logistic regression, and Chi-squared tests (or Fisher's exact test for cell counts less than five) will be reported.

**Section D: Multiple imputation process and software**

The primary analysis will assume that missing data are Missing at Random (MAR) and will be handled using Multiple Imputation (MI), rather than complete case analysis. Although complete case analysis provides unbiased estimates under the more restrictive Missing Completely at Random (MCAR) assumption, MI is valid under both MAR and MCAR, and increases statistical power by utilising information from all available observations.

Imputation will be performed using the *mice* package in R (Multiple Imputation by Chained Equations). The imputation model will include all available covariates including treatment assignment, hospital and all outcome variables. A minimum of m = 20 imputed datasets will be generated, and results will be combined using Rubin's Rules to obtain the final estimates.

**Section E: Handling and analysis of censored data**

For the time-based variables (LOS, time to clear fluids, free fluids, full feeds, passage of stool), censored observations represent incomplete information rather than missing data and will be handled in the survival models. Our analyses rely on the Censoring at Random (CAR) assumption, which assumes that the timing of censoring does not provide additional information about the survival outcome or event times, conditional on the covariates included in the model. Therefore, any baseline covariates believed to be associated with the censoring process will be included as covariates in the RMST and Cox proportional hazards models. If there is a strong *a priori* concern that censoring may be informative such that the CAR assumption is violated, we will conduct a sensitivity analysis to assess the robustness of our conclusions. Potential methods for this analysis may include a competing risks analysis such as a the Fine-Gray model, depending on the nature of the censoring events.
