## Supplementary material for "Early enteral nutrition after Paediatric Ostomy Closure (EPOC): a protocol for a multicentre, prospective randomised controlled trial": Online Supplementary Appendix 5

Study Number:______

#### Patient Data Collection Booklet

**CONFIDENTIAL**
This document is confidential and the property of Sydney Children's Hospital. No part of it may be transmitted, reproduced, published, or used without prior written authorization from the institution.

**Statement of Compliance**
This document contains patient information for the clinical trial. This study will be conducted in compliance with all stipulation of this protocol, the conditions of the ethics committee approval, the NHMRC National Statement on ethical Conduct in Human Research (2007) and the Note for Guidance on Good Clinical Practice (CPMP/ICH-135/95).

### Parent / Guardian Consent Form

| | |
|---|---|
| **Study Title** | Early enteral nutrition after Paediatric Ostomy Closure (EPOC): A Prospective Randomised Controlled Trial |
| **Principal Investigator/s** | Susan Adams - Department of Paediatric Surgery, Sydney Children's Hospital, Randwick. 9382 1776<br>Soundappan Soundappan - Department of Paediatric Surgery, Children's Hospital at Westmead, 9845 3235 |
| **Main Study Contact Person** | Susan Adams - 9382 1776 |

*Declaration by Parent / Guardian*

☐ I have read the Parent / Guardian Information Sheet or someone has read it to me in a language that I understand.

☐ I understand the purposes, procedures and risks of the research project described in the Parent / Guardian Information Sheet.

☐ I have had an opportunity to ask questions and I am satisfied with the answers I have received.

☐ I freely agree to the child participating in this research project as described and understand that I am free to withdraw them at any time during the project without affecting their future health care.

☐ I understand that I will be given a signed copy of this document to keep.

☐ I give permission for my child's treating doctor, other health professionals, hospitals or laboratories outside this hospital to release information to UNSW concerning my child's condition and treatment for the purposes of this project. I understand that such information will remain private and confidential.

☐ I understand that the results of this project, may be given for the use of data in future research projects that are: an extension of the original project; or in the same general area of research.

☐ I wish to receive a lay summary of the study findings via the following email / post address:

____________________________________________________________

Name of Child (if applicable): ____________________________________________

Signature of Child (if applicable): ________________________ Date: ________________

Name of Parent / Guardian (please print): ________________________________________

Signature of Parent / Guardian: ______________________ Date: _________________

*Under certain circumstances (see Note for Guidance on Good Clinical Practice CPMP/ICH/135/95 at 4.8.9) a witness* to informed consent is required.*

Name of Witness* to Parent / Guardian's Signature (please print): __________________________

Signature of Witness: _______________________________ Date: __________________

* The Witness is not to be the investigator, a member of the study team or their delegate. In the event that an interpreter is used, the interpreter may not act as a witness to the consent process. Witnesses must be over 18 years of age

| | **YES** | **NO** |
|---|---|---|
| Consent scanned into REDCap | | |

the children's hospital at Westmead + SYDNEY CHILDREN'S HOSPITAL RANDWICK = The Sydney Children's Hospitals Network (Randwick and Westmead)

#### Eligibility criteria

| Inclusion criteria: (should be ticked yes) | YES | NO |
|---|---|---|
| Between 3 months and 16 years | | |
| Elective enterostomy | | |
| Closure of colostomy after ARM repair **OR** | | |
| Closure of ileostomy after previous neonatal laparotomy for NEC, perforation, atresia, or other intestinal pathology **OR** | | |
| Closure of colostomy/ileostomy where the stoma was formed at a previous laparotomy for other intraabdominal pathology | | |

| Exclusion criteria: (should be ticked no) | YES | NO |
|---|---|---|
| Patients with short gut requiring long term total parenteral nutrition (TPN) | | |
| Patients undergoing a total colectomy or gastrostomy closure | | |
| Intercurrent conditions expected to affect bowel function (e.g., neurological impairment) | | |
| Patients who have been assessed to have unsafe swallow or laryngeal penetration with reflux and vomiting. This includes inability to protect the airway due to neurological impairment or structural abnormality, due to aspiration risk. | | |
| Inability to obtain consent from parent/guardian | | |
| Patients requiring specific individualised feeding regime (e.g., metabolic disease) | | |
| Patients who require prolonged nasogastric tube (NGT) feeding or gastrostomy feeding prior to and following surgery, that do not have normal gut function | | |
| Patients requiring more than a simple enterostomy closure; i.e., extensive adhesiolysis or more than one anastomosis (in NEC) | | |
| Patients with Cystic Fibrosis or Inflammatory Bowel Disease. | | |

**If YES ticked for any of the exclusion criteria – do not include the child in the study.**

the children's hospital at Westmead + SYDNEY CHILDREN'S HOSPITAL RANDWICK = The Sydney Children's Hospitals Network (Randwick and Westmead)

#### Randomisation data

**Date of Randomisation:** ☐☐ ☐☐ ☐☐☐☐

**Date of Birth:** ☐☐ ☐☐ ☐☐☐☐

**Medical Record Number:** ______________________________

**Study ID:** ☐☐☐

**<u>Strata information</u>**

**Hospital Location**

☐ SCH ☐ CHW

**Age Group**

☐ 3m to <6y ☐ 6y to <16y

**Stoma Location**

☐ Small bowel ☐ Large bowel

**<u>Randomisation slip:</u>** (paste below)

#### Other admission data

**Operation time:** ______________________________

**Admission weight:** ______________________________

**Discharge weight:** ______________________________

**Admission height:** ______________________________

**Discharge weight:** ______________________________

**Pain scores:**

| Date | Score | Comments |
|---|---|---|

the children's hospital at Westmead + SYDNEY CHILDREN'S HOSPITAL RANDWICK = The Sydney Children's Hospitals Network (Randwick and Westmead)

If breast fed: for volume put time of feed (or small, medium or large feed)

### Bedside data sheet

#### Day 1 Date:

| Time | Intake | | Output | | Details |
|---|---|---|---|---|---|
| | Type of intake | Volume | Stool | Vomit | |

**Day 2** **Date:**

| Time | Intake | | Output | | Details |
|---|---|---|---|---|---|
| | Type of intake | Volume | Stool | Vomit | |

the children's hospital at Westmead + SYDNEY CHILDREN'S HOSPITAL RANDWICK = The Sydney Children's Hospitals Network (Randwick and Westmead)

**Day 3** **Date:**

| Time | Intake | | Output | | Details |
|---|---|---|---|---|---|
| | Type of intake | Volume | Stool | Vomit | |

the children's hospital at Westmead + SYDNEY CHILDREN'S HOSPITAL RANDWICK = The Sydney Children's Hospitals Network (Randwick and Westmead)

#### Day 4 Date:

| Time | Intake | | Output | | Details |
|---|---|---|---|---|---|
| | Type of intake | Volume | Stool | Vomit | |

the children's hospital at Westmead + SYDNEY CHILDREN'S HOSPITAL RANDWICK = The Sydney Children's Hospitals Network (Randwick and Westmead)

**Day 5** **Date:**

| Time | Intake | | Output | | Details |
|---|---|---|---|---|---|
| | Type of intake | Volume | Stool | Vomit | |

**Day 6** **Date:**

| Time | Intake | | Output | | Details |
|---|---|---|---|---|---|
| | Type of intake | Volume | Stool | Vomit | |

#### Parent satisfaction form

Please answer the following questions telling us how happy you are with the care you, your child, and your family have received at the hospital and after discharge from hospital from the staff. Please tell us how happy you are with each item by circling:

***Please add any further comments about your child's care during the trial and their hospital stay***

Thank you for your participation!

#### Protocol violations

| Any protocol violation occurred (tick one): | YES |
|---|---|
| No protocol violations | |
| One protocol violation | |
| Two protocol violations | |
| Three protocol violations | |
| Four or more protocol violations *How many?* | |

**Details of first protocol violation**

______________________________________________

______________________________________________

**Time of first protocol violation (in 24 hour time):** | | | | |

**Reason for first protocol violation**

______________________________________________

______________________________________________

**Details of any second protocol violation**

______________________________________________

______________________________________________

**Time of any second protocol violation (in 24 hour time):** | | | | |

**Reason for any second protocol violation**

______________________________________________

______________________________________________

**Details of any other protocol violations**

______________________________________________

______________________________________________

______________________________________________
